# Epithelial brushing storage conditions and the reliability of IF staining – implications for primary ciliary dyskinesia diagnostics

**DOI:** 10.1101/2025.07.01.25330648

**Authors:** Przystalowska-Maciola Hanna, Dabrowska Malgorzata, Zietkiewicz Ewa, Bukowy-Bieryllo Zuzannna

## Abstract

Immunofluorescence (IF) microscopy is gaining increased popularity as a pre-genetic diagnostic method in primary ciliary dyskinesia (PCD). Efficient IF-based diagnostics in PCD requires standardization of staining methods and antibodies. Obviously, specificity of the analysis highly depends on the quality of the analyzed material. The nasal epithelium brushing quality depends on the patient health status, but conditions of the slides storage are also very important.

We applied automated image analysis to systematically examine the influence of samples’ storage conditions on the specificity of IF staining with eight polyclonal antibodies routinely used for PCD diagnostics, against epitopes of proteins representing various axoneme structural complexes. Seven various combinations of temperatures and storage times were tested to mimic the procedures of handling epithelial brushing on glass slides: at the clinic, during transport, or after reception at the diagnostic laboratory.

Our study revealed that proper slide storage conditions are essential for the reliable PCD diagnosis via IF staining. If microscopic slides are prepared at the diagnostic laboratory, we suggest continuous storage at -80°C or -20°C. If the slides are prepared at a collaborating clinic and shipped, we suggest storage at -20°C or 4°C. The IF sensitivity to slide storage conditions differs among antibodies targeting various ciliary elements; for example, molecular ruler proteins appear the most sensitive to prolonged slide storage at room temperature. Moreover, to improve the reliability of the IF staining, additional control slides should be used to account for inter-individual differences. Finally, IF results indicative of PCD should be carefully confronted with patient’s clinical history.

## Introduction

The majority of human respiratory tract is lined with the specialized airway epithelium, composed of several cell types, including multiciliated cells, which carry hundreds of motile cilia on their apical surface [1]. Motile cilia, whose main role is to transport fluid and mucus in the organism, are microtubule-based organelles, composed of the axoneme anchored in the cell membrane by the basal body [2]. The ciliary axoneme is composed of 9 microtubular (MT) doublets surrounding a central pair (CP) complex. Peripheral doublets are connected to the central pair complex by radial spokes (RS), and between each other by nexin-dynein regulatory complexes (N-DRC). Peripheral MT doublets of the motile cilia are decorated through the axoneme length with multiprotein complexes, including inner (IDA) and outer (ODA) dynein arms responsible for generating the ciliary movement, and “molecular ruler” (MR) responsible for proper spacing between dynein arms along the axoneme [3].

Genetically determined defects of the motile cilia function (resulting from changes in the structure or number of these organelles) lead to a rare multisystemic disease – primary ciliary dyskinesia (PCD), with the occurrence estimated between 1:10,000 and 1: 20,000 [4, 5]. The main symptoms of PCD include chronic upper and lower respiratory tracts infections, abnormal mucociliary clearance, recurrent otitis media, infertility, and in 50% of cases randomization of organ laterality [6]. Genetically, PCD is highly heterogeneous. It is most frequently inherited in an autosomal recessive, and rarely in the X-chromosomal or autosomal dominant manner. Hundreds of pathogenic variants have been so far identified in over 50 different genes [3, 7]. In spite of significant developments in the field of PCD genetics, successful detection of underlying defects is presently achieved in only 60-70% of patients [8].

Due to unspecific symptoms, PCD diagnosis is often delayed and complicated [9]. The first-line diagnostic tests in PCD, which precede identification of pathogenic genetic variants, include: (1) nasal nitric oxide (nNO) measurement, (2) ciliary motility assessment using high-speed videomicroscopy analysis (HSVMA), (3) analysis of motile cilia structure using transmission electron microscopy (TEM), (4) immunofluorescent (IF) microscopy of the respiratory epithelium cells allowing visualization of ciliary proteins [2, 10].

IF microscopy is gaining increased popularity as a pre-genetic diagnostic method in PCD [11–14]. By revealing the absence or mislocalization of specific proteins – markers of the ciliary ultrastructure elements – it can confirm axonemal defect in the material collected from a patient [15]. While not always indicating the defective protein itself, it can be used to guide genetic testing rendering it more specific and cost-effective [16]. An important practical advantage of the IF-based analysis is that collecting samples (typically, nasal epithelium brushing spread and dried on glass microscope slides, further referred to as slides) can be done by medical personnel with no previous experience in PCD, and shipped to specialized diagnostic laboratories [14].

Efficient and reliable IF-based diagnostics in PCD requires standardization of staining methods and antibodies. One of the challenges in the IF-based diagnostics concerns detection of genetically determined defects in the ciliary protein synthesis or assembly, which result in weakening of the IF signal in cilia, accompanied by the presence of non-specific diffused signal in the cytoplasm. We hypothesized that similar patterns may be observed when the quality of analyzed material is compromised. Obviously, the nasal epithelium brushing quality depends on the donor’s health status (inflammation and infection can obscure the diagnosis [16]), but conditions of the slides storage are also very important. So far, no study has been performed to quantify and formally test the influence of slide storage conditions on the specificity of IF staining.

The aim of the study was to compare reliability of IF staining with selected polyclonal antibodies routinely used for PCD diagnostics at a range of slide storage conditions, chosen to reflect realistic scenarios of slides handling between sample collection and the IF analysis. The results of the study provide clues concerning proper slides’ handling, adherence to which should improve reliability of the IF-based PCD diagnostics.

## Results

Slides with nasal brushings from five healthy donors were stored at seven time and temperature combinations **(Table 1)**. Storage at -80°C for 28 days was assumed to represent best storage conditions. The effect of storage at an alternative temperature of -20°C was tested for 28 days or 8 weeks. To mimic shipping of slides between clinics and diagnostic laboratories, three more conditions were examined: -20°C, 4°C or RT for 11 days (clinics) followed by 3 days at RT (transport), and 14 days at -80°C (lab). Slides stored for 28 days at RT were considered detrimental (the most unfavorable of the tested storage conditions).

**Table 1.**
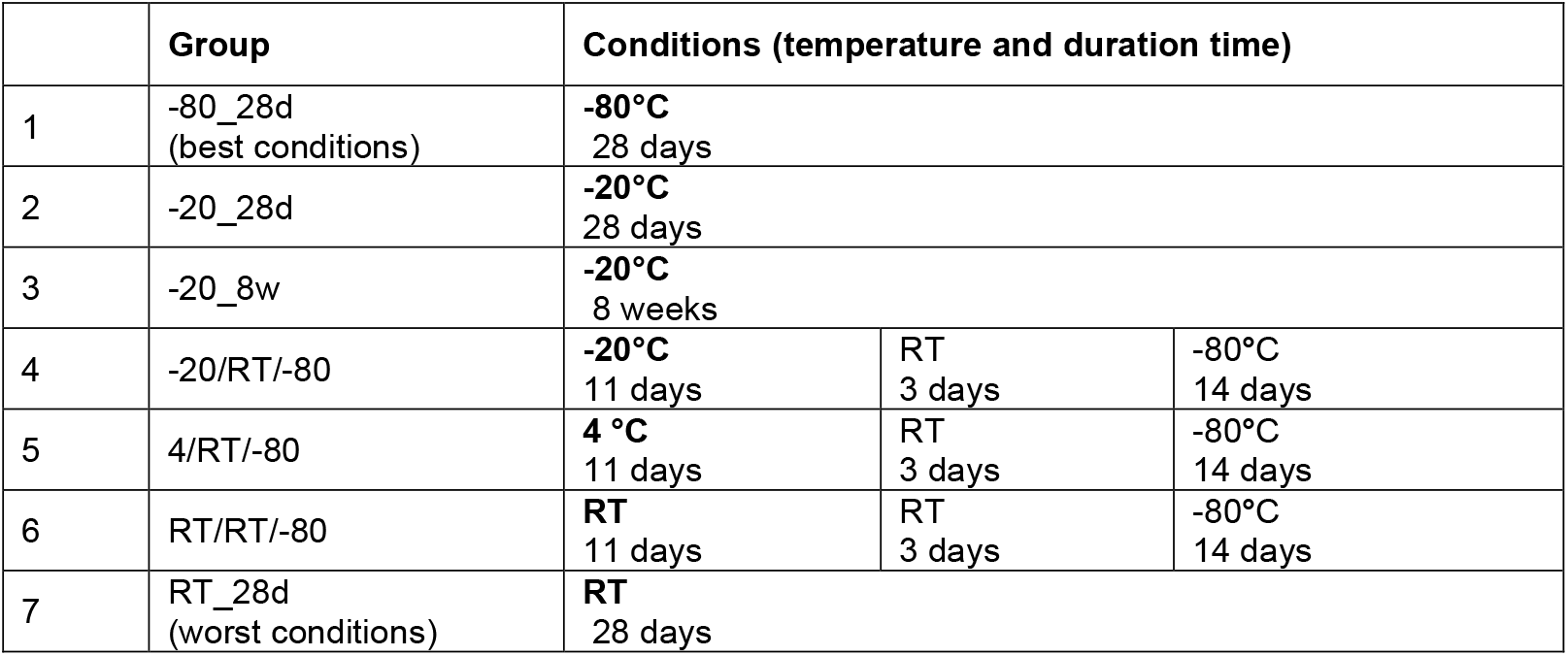
Storage conditions.

After the end of the storage period, slides were stained with eight polyclonal rabbit antibodies commonly used in PCD diagnostics (see **Table S1);** the secondary anti-rabbit IgG antibody staining was visualized in the red channel. Antibodies were selected to target proteins representing various elements of the axoneme, and throughout the text are referred to using targeted protein names (DNAH5 – ODA, DNALI1 – IDA, RSPH4A and RSPH9 – RS, GAS8 – N-DRC, CCDC39 and CCDC40 – MR, SPEF2 – CP complex).

Each slide was also treated with a monoclonal mouse antibody against acetylated α-tubulin (Ac-αTub), a component of the ciliary MT; the secondary anti-mouse IgG antibody staining was observed in the green channel. The Ac-αTub staining was clearly visible and specific across the analyzed conditions, and served as a stable marker of the ciliary axoneme. An overlap of the red signal with the green marker indicated the presence of the target protein in cilia.

The specificity of staining using tested polyclonal antibodies was highly variable depending on the storage time and temperature; the size of the “storage effect” depended on the targeted ciliary proteins. Representative images of the IF staining of slides stored at the positive control versus the most detrimental conditions (negative control) are shown in **Figure 1**.

**Figure 1.**
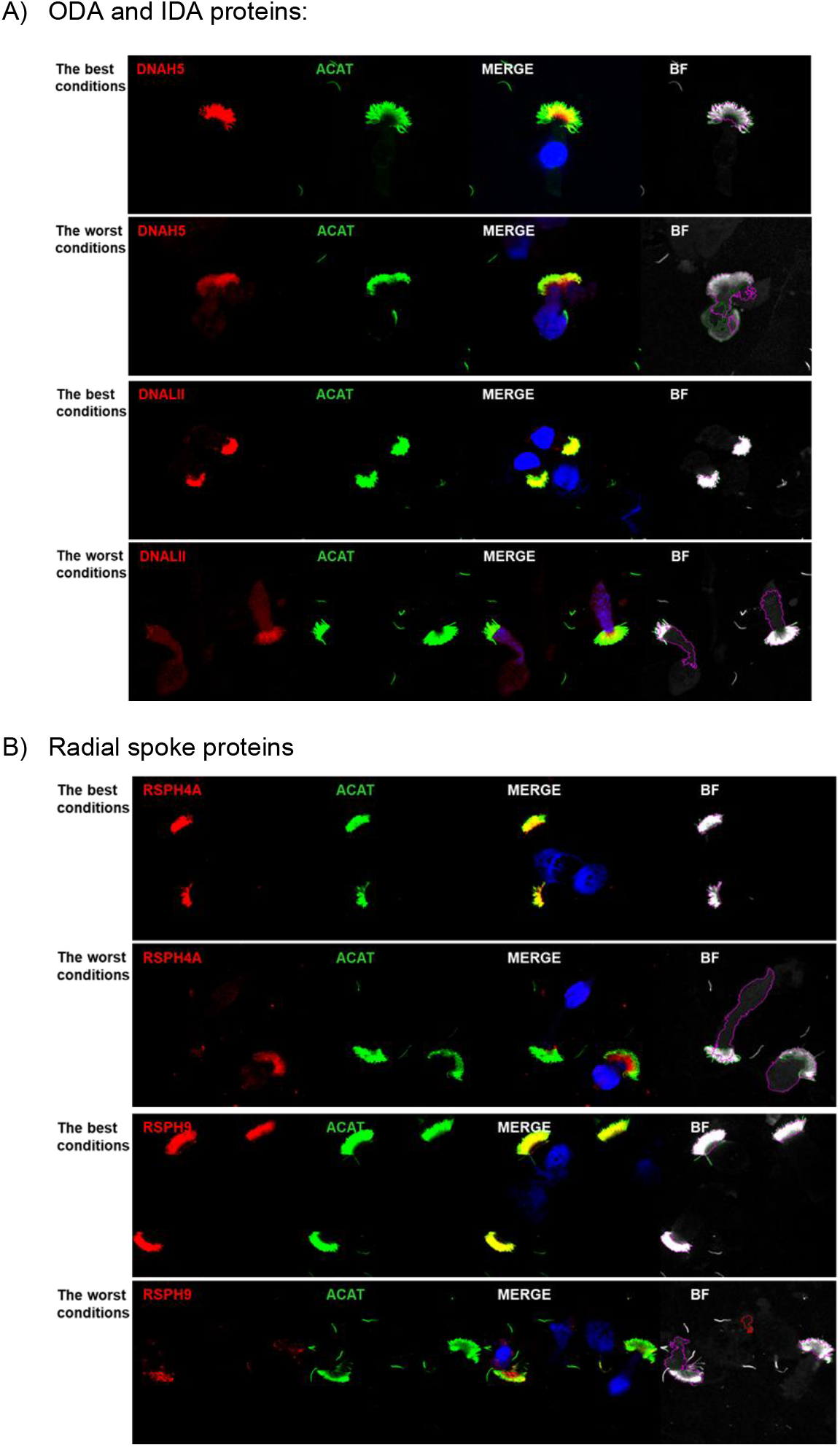
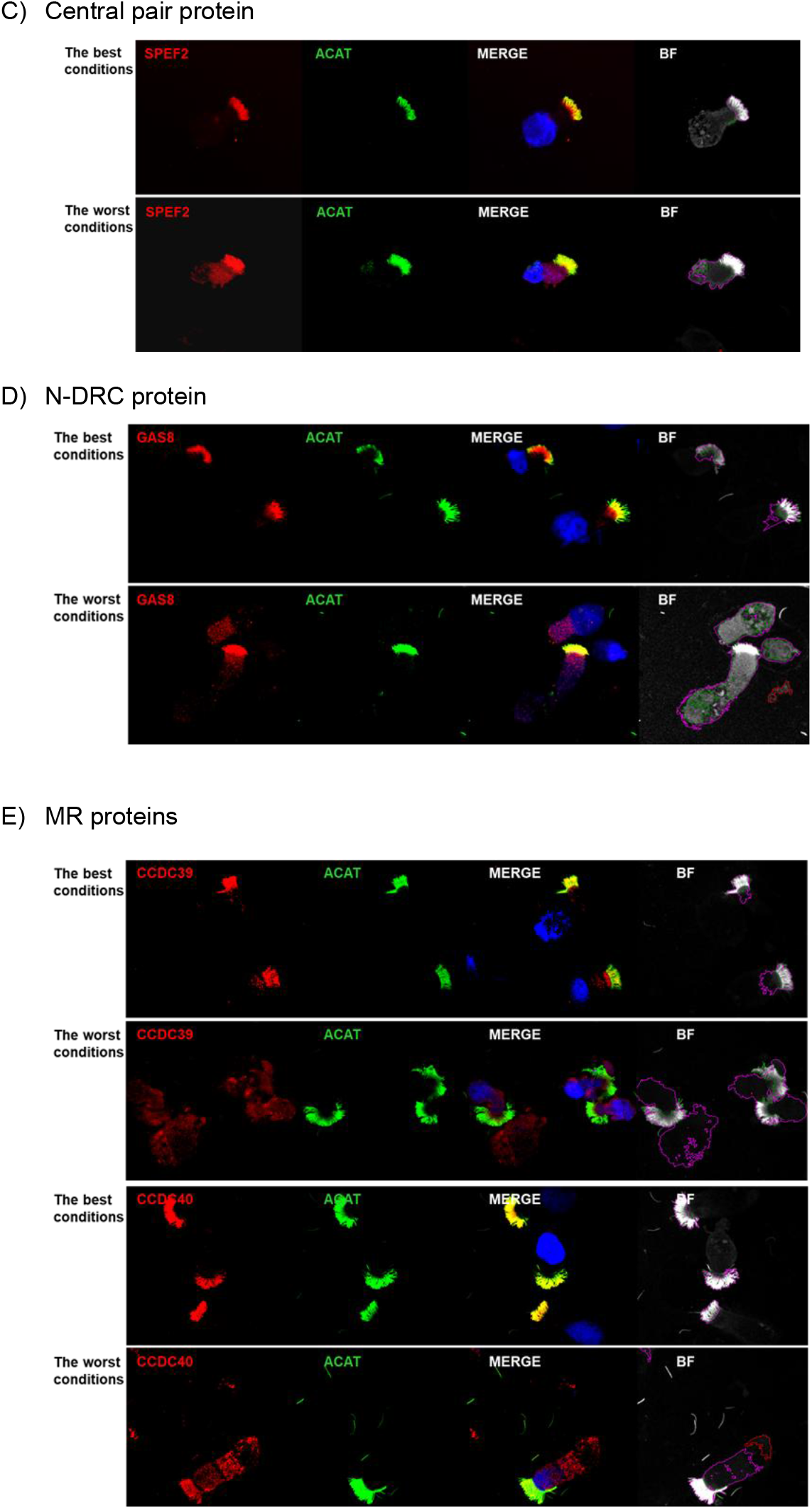
Representative IF images of various target proteins distribution (red) and the axoneme marker (green), at border storage conditions (−80°C/28d: the best conditions, versus RT/28d: the worst conditions). (A) ODA and IDA proteins; (B) RS proteins; (C) CP complex protein; (D) N-DRC protein; (E) MR proteins. Nuclei were stained with DAPI (blue).

### Image analyses using CellProfiler

In order to provide quantitative measure of the slide storage conditions’ impact on the IF staining, automated image analysis was performed using a custom Cell Profiler pipeline **(Figure 2)**.

**Figure 2.**
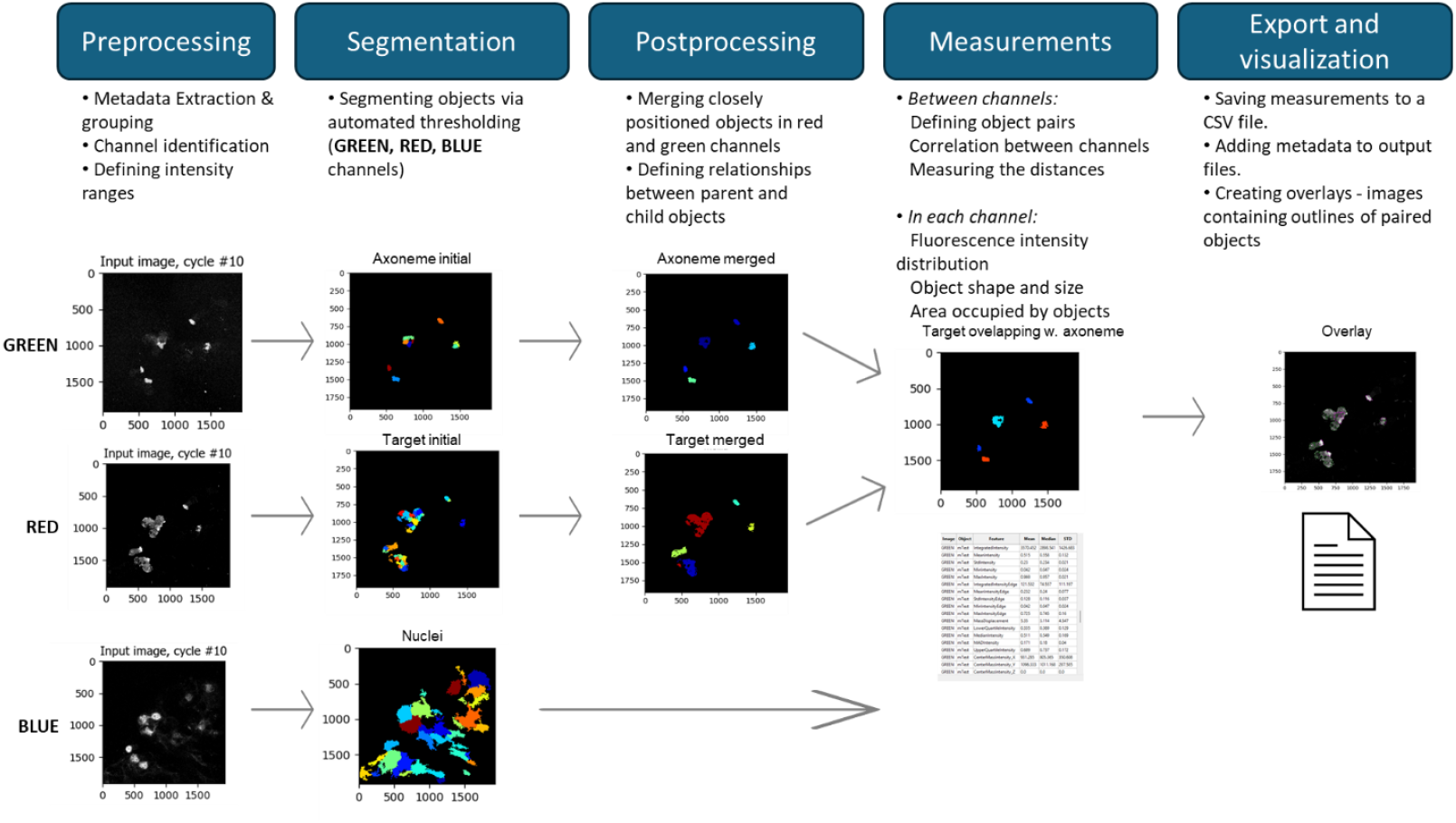
Custom Cell Profiler pipeline.

In total, 280 slides from 5 donors across 56 combinations of storage conditions (7) and antibodies (8) were analyzed. For each combination, the values of parameters analyzed in CellProfiler were presented as the means of individual results obtained for five tested donors. In total, 6167 images, typically representing 1-5 cells, were analyzed (on average, 22 images per one slide; more detailed information below and in the **Supplementary file**).

Image analysis in the CellProfiler pipeline returned two main types of parameters characterizing objects in each channel (objects’ size, shape, and fluorescence intensity distribution) and characterizing the relations between the channels (general channel correlation and the distance between object pairs from the same cell, present in red and green channels). All the parameters confirmed unchanged stability of the green objects (Ac-αTub staining) across various storage conditions; the green color (marker of the axoneme) was therefore used only as a reference.

Among 114 parameters analyzed in CellProfiler, 81 significantly correlated with slide storage conditions (p < 0.001, **Supplementary Table S2**). The majority of them characterized red channel objects in terms of their area & shape, and intensity (29 and 43 parameters, respectively). The second type of parameters described the red-versus-green channel comparisons, preceded by the definition of paired objects from both channels (seven of them estimated correlation between the channels, and one measured distance between the centroids of paired objects) (**Supplementary Table S2**). For further presentation, representative parameters from each parameter group were chosen. For each of the selected parameters, a heat map was prepared, showing the mean value of the parameter (averaged for slides from five individuals) for 56 combinations of storage condition and tested antibody. To assure that the analyzed objects from various channels were present in one cell, only the paired objects were taken for the parameter analysis.

### Channel parameters

#### Area characteristics

The impact of slide storage conditions on the area, eccentricity and compactness of the objects’ in the red channel was compared for various tested antibodies (**Figure 3**). At a given storage conditions, the parameters’ values indicating cilia-specific staining using a tested antibody were most similar to those characterizing a stable axoneme marker: low object’s area and low compactness (indicating shape regularity), as well as high eccentricity (range 0-1, higher values indicating stretched rather than circular shape).

**Figure 3.**
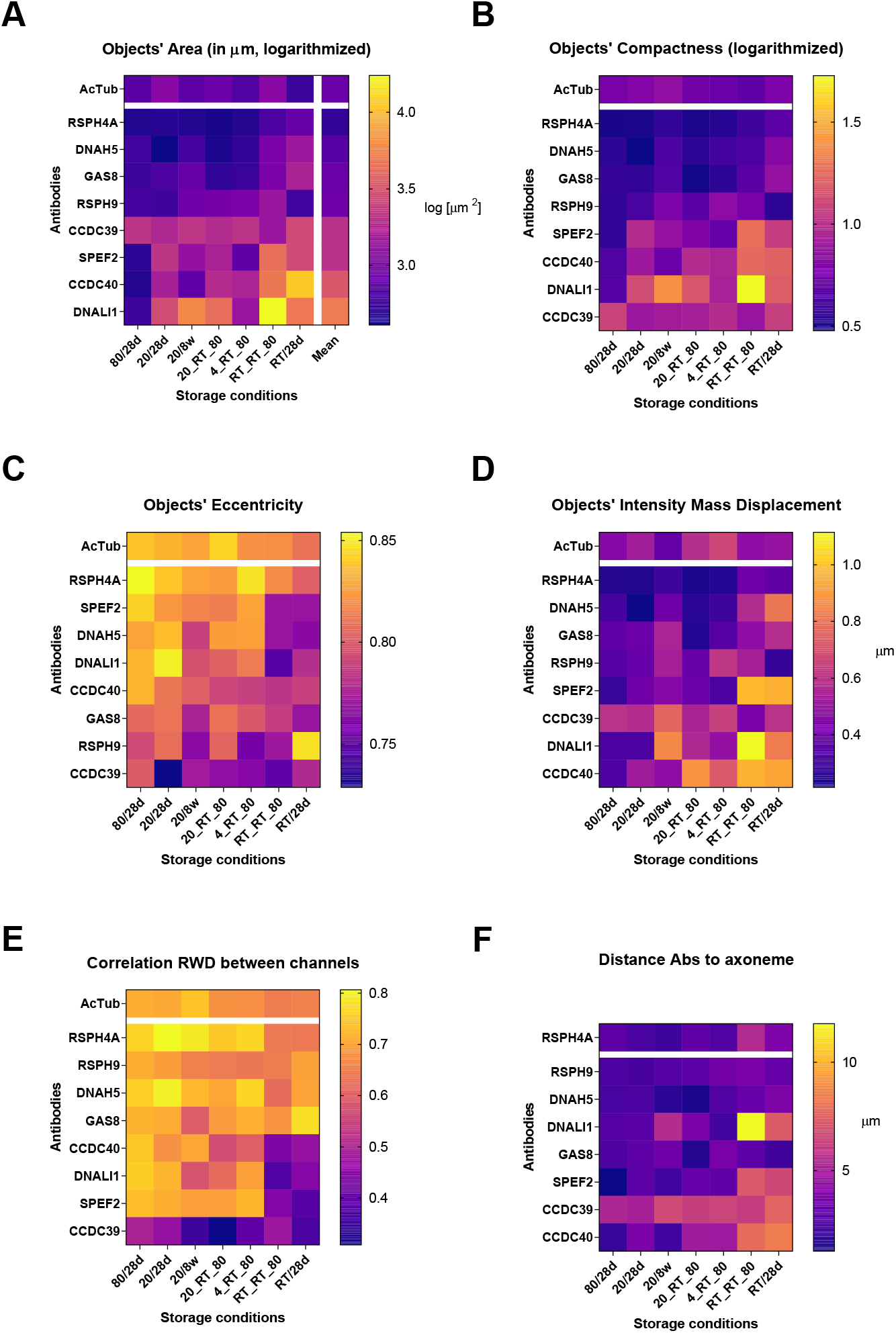
Comparison of the red objects’ characteristics. (A) Area; (B) Compactness; (C) Eccentricity; (D) Intensity Mass Displacement; (E) Correlation RWD, (F) Distance to axoneme marker. The parameters (values represented by colors in the heat map) were calculated separately for each tested antibody and storage condition. Heatmap legends are ordered according to the parameter’s values indicating highest compatibility with the axoneme marker; the order of antibodies is according to their averaged parameter value across all conditions (see column to the right of each heatmap). For graphs in A, B values have been logarithmized to improve the heatmaps’ utility and facilitate interpretation. The order of storage conditions (on X axes) is identical in A-F.

For antibodies targeting RSPH4A, RSPH9, DNAH5 or GAS8, the area size and compactness values were low (similar to axonemal marker) across various slide storage conditions (**Figure 3A, 3B**). Antibodies targeting CCDC39, CCDC40, SPEF2, and DNALI1 displayed lower specificity of staining (higher object area and compactness) for slides stored at less favorable conditions, especially those including prolonged storage at RT. Eccentricity values yielded less consistent results. High values (similar to the axoneme marker, indicating high staining specificity) were observed for RSPH4A across various conditions, even those including prolonged times at RT that were detrimental to staining with other antibodies (**Figure 3C**). Surprisingly, prolonged slide storage at -20°C resulted in low eccentricity values for staining with RSPH9, GAS8 and DNAH5.

#### Intensity parameters

Intensity MassDisplacement parameter measures the distance between the intensity-weighted center of the object and its geometric center: the higher the distance, the more asymmetric is the signal intensity distribution within the object. The calculated values indicated various levels of red signal distribution within the cell **(Figure 3D)**. Compared to other conditions, prolonged storage at RT (RT_28d, RT/RT/-80) resulted in a marked increase of this parameter, suggesting that the red signal was broadly distributed within the cell without accumulation in cilia (**Figure 3D**). This is in agreement with the IF images, where storage at RT_28d, compared to -80_28d, caused visible reduction in the red staining within the cilium, and increase in the red staining in the cytoplasm (**Figure 1**). At storage conditions, which did not involve prolonged incubation at RT, the lowest red signal asymmetry was observed for RSPH4A, RSPH9, DNAH5, GAS8 proteins, while for SPEF2, DNALI1 and CCDC39, CCDC40 the asymmetry was strikingly higher.

#### Analysis between channels

##### Pairing of tested antibodies with the axoneme marker

Among 6167 analyzed images, Cell Profiler pipeline detected 31566 objects in the red channel, of which 23347 (∼74%) at least partially overlapped with objects identified in the green channel (axoneme marker). Note that the percentage of unpaired objects may include, both, unpaired red and green objects within the same cell, and objects belonging to different cells.

##### Correlation characteristics

**RED_GREEN** Correlation, with values ranging from -1 to 1, is an image-based (as opposed to object-based) parameter, which assesses how changes in the red signal intensity are linked to changes in the green signal at the same spatial locations within the image. High positive values indicate that when the red is high (or low), the signal in the green channel tends to be high (or low) as well; values lower than 0 suggest a reverse correlation. The RED-GREEN correlation was at the top of the list of the parameters significantly correlating with storage conditions (it had the smallest p-value of 2.30E-50) **(Supplementary Table S2)**. Of note, the GREEN-RED Correlation parameter (describing correlation between the localization and intensity of the green signal with that of the red signal) was not significantly associated with storage conditions (p-value 0.75)(**Supplementary Table S2)**.

Analysis of the overall RED_GREEN correlation values (averaged across all tested antibodies) confirmed that storage at RT for longer times (RT/RT/-80 and RT_28d) resulted in a considerable reduction of this parameter (**Figure 3E**).

Apart from these longer RT storage conditions, the RED-GREEN correlation values were usually rather stable for antibodies targeting RS, ODA and CP. Antibodies targeting IDA and MR proteins displayed less consistent results; the lowest RED-GREEN correlation values across different storage conditions were observed for CCDC39 (**Figure 3E**).

##### Distance between the object pairs

Distance_Centroid_mCilia parameter measures the distance between the geometric centers (centroids) of the objects in the red channel (tested antibodies) and green channel (axoneme marker). The increase in the area and shape differences of the objects in the analyzed channels, observed under different storage conditions, was also reflected in considerable differences in the Distance_Centroid_mCilia.

Heatmap comparison of the distances for each tested antibody and storage conditions is shown in **Figure 3F**. For CCDC39, even at the “positive control” conditions, the average distances between the signal and axoneme marker were significantly higher (6.07 ±0.73 µm vs 3.56 ± 0.83 µm, p-value <0.0001) than for all other tested antibodies (**Figure 3F**).

##### Integrated Performance Metrics

The ranks obtained for each of the analyzed parameters were averaged into one metric, which assessed the most optimal storage conditions across all tested antibodies (**Table 2**) and the general performance of each tested antibody averaged across the storage conditions (**Table 3**).

**Table 2.** General performance ranks of slide storage conditions. The numbers in the table correspond to the order of conditions in the heatmap (1 being best, 7 being worst). For each of the analyzed parameters, slide storage conditions were ranked according to their overall performance, and reflects the conditions’ order in each heatmap. This overall performance rank was calculated as the mean from all parameters, colors of the bars below the table indicate the performance (green – very good, yellow – average, red-poor)

| Storage condition | -80_28d | -20_28d | -20/RT/-80 | -20_8w | 4/RT/-80 | RT/RT/-80 | RT_28d |
| --- | --- | --- | --- | --- | --- | --- | --- |
| Mean_mTest_Correlation_RWC_RED_GREEN | 1 | 2 | 5 | 4 | 3 | 6 | 7 |
| Mean_mTest_AreaShape_Compactness | 1 | 2 | 3 | 4 | 5 | 6 | 7 |
| Mean_mTest_AreaShape_Area | 1 | 2 | 4 | 3 | 5 | 6 | 7 |
| Mean_mTest_Area_Eccentricity | 1 | 2 | 3 | 5 | 4 | 6 | 7 |
| Mean_mTest_Intensity_Mass_Displacement | 1 | 3 | 2 | 5 | 4 | 7 | 6 |
| Mean_mTest_Distance_Centroid_mCilia | 1 | 2 | 4 | 3 | 5 | 7 | 6 |
| Overall Performance Rank: | 1,0 | 2,2 | 3,5 | 4,0 | 4,3 | 6,3 | 6,7 |

**Table 3.** Overall performance of analyzed antibodies. For each of the analyzed parameters, antibodies were ranked according to the mean parameter’s value for all storage conditions (see rightmost columns in the heatmaps). The overall performance rank was calculated as the mean from all parameters. Colors of the bars below the table indicate the performance (green – very good, yellow – average, red – poor). Calculations were made for green objects pairing with only 1 red object.

|  | RSPH4A | DNAH5 | GAS8 | RSPH9 | SPEF2 | CCDC40 | DNALI1 | CCDC39 |
| --- | --- | --- | --- | --- | --- | --- | --- | --- |
| Mean_mTest_Correlation_RWC_RED_GREEN | 1 | 2 | 3 | 4 | 5 | 6 | 7 | 8 |
| Mean_mTest_AreaShape_Compactness | 1 | 2 | 3 | 4 | 5 | 6 | 7 | 8 |
| Mean_mTest_AreaShape_Aarea | 1 | 2 | 3 | 4 | 5 | 6 | 7 | 8 |
| Mean_mTest_Area_Eccentricity | 3 | 6 | 4 | 2 | 1 | 7 | 5 | 8 |
| Mean_mTest_Intensity_Mass_Displacement | 1 | 2 | 3 | 4 | 5 | 8 | 7 | 6 |
| Mean_mTest_Distance_Centroid_mCilia | 4 | 1 | 2 | 3 | 5 | 6 | 7 | 8 |
| Overall Performance Rank: | 1,8 | 2,5 | 3,0 | 3,5 | 4,3 | 6,5 | 6,7 | 7,7 |

The best storage conditions (**Table 2**, green) were -80_28d and -20_28d. Increased time of storage at

-20°C caused a slight drop in the performance of some of the antibodies. Storage conditions including the short-term storage at RT (“shipping”) were less favorable, although the RT incubation only for 3-4 days did not considerably influence IF results (**Table 2**, yellow). The two definitively worst storage conditions were those including a long-term storage at RT(RT_28d, RT/RT/-80) (**Table 2**, red).

Three best performing antibodies were those targeting RSPH4A, DNAH5 and GAS8, while antibodies against MR proteins and DNALI1 were characterized by the worst performance (**Figure 3; Table 3**). The poor performance of the latter antibodies appeared to be caused by their large sensitivity to long storage at RT (**Figure 3**). However, even at storage conditions excluding prolonged RT, they performed worse than other antibodies (**Table 4, Supplementary Table S3**). Interestingly, GAS8, which was relatively insensitive to the prolonged RT, performed worse than RSPH4A and DNAH5 at other conditions.

**Table 4.** Comparison of the antibody performance ranks obtained for all storage conditions and storage conditions excluding prolonged RT storage. Colors of the fields indicate the antibody performance (green – very good, yellow – average, red – poor). Calculations were made for green objects pairing with only 1 red object.

| Antibody | All storage conditions | All conditions except prolonged RT |
| --- | --- | --- |
| RSPH4A | 1.8 | 1.5 |
| DNAH5 | 2.5 | 2.2 |
| GAS8 | 3.0 | 4.2 |
| RSPH9 | 3.5 | 4.5 |
| SPEF2 | 4.3 | 4.2 |
| CCDC40 | 6.5 | 5.7 |
| DNALI1 | 6.7 | 6.0 |
| CCDC39 | 7.7 | 7.8 |

## Discussion

We applied automated image analysis to systematically examine the influence of slides storage conditions on the specificity of IF staining with antibodies routinely used for PCD diagnostics. In an attempt to identify best storage conditions applicable in the clinical practice, we have tested various combinations of temperatures and storage times, which mimic the procedures of handling epithelial brushing on glass slides: at the clinic, during transport, or after reception at a diagnostic laboratory.

### Slide storage condition

The analysis of slide storage conditions indicated that both storage temperature and time were important factors influencing IF staining specificity.

As expected, the long-term (>14 days) storage at RT had the worst impact on the IF staining specificity, as evidenced by the lowest overall performance scores (**Table 2**). For the majority of parameters tested, four weeks of storage at RT, or two weeks of storage at RT followed by 2 weeks at -80°C yielded the worst values of tested parameters (**Figures 1, 3, Table 2**).

Uninterrupted 4-week storage at -80°C, or at -20°C, had the least impact on the staining by tested antibodies; prolongation of the storage at -20°C to 8 weeks did not lead to worse staining results.

Comparison of the results for conditions containing an intermittent RT incubation period, mimicking transport of slides, indicates that the short-term increase of the temperature had no remarkable effect on the parameters describing the specificity of IF staining (**Figure 3**).

Our results indicate that samples to be shipped from the clinics to diagnostic laboratories should be stored at -20°C or at 4°C; as samples are not affected by a short incubation at RT, the shipment can be done at

RT. After reception at a diagnostic laboratory, samples should be stored at -80°C until IF analysis is performed (**Figure 3, Table 2**).

### Epitope sensitivity to storage conditions

Specific IF detection of various ciliary complexes using antibodies targeting RS (RSPH4A, RSPH9), ODA (DNAH5), N-DRC (GAS8), and CP protein (SPEF2) was relatively insensitive to various slide storage conditions, in case of RSPH9 and GAS8 including long-term exposure to the RT. Staining specificity of IDA protein (DNALI1) staining was very vulnerable to the prolonged slide storage at RT, but even more so to repeated freeze-thawing process. CCDC39 detection was the most sensitive to storage conditions, with satisfactory results observed only when slides were stored at -80oC (**Figures 1**,**3, Tables 3, 4**).

IF detection of both MR proteins (CCDC39 and CCDC40) (**Tables 3, 4, Supplementary Table S3**) was negatively affected by almost all slide storage conditions, rendering CCDC39 or CCDC40 antibodies unsuitable in IF based-search for MR defects. Fortunately, these defects have been previously shown detectable by IF staining using GAS8 antibody [12] [17–19], which in our study was storage condition-resistant **(Tables 3, 4, Supplementary Table S3)**. If using the CCDC39 antibody is for some reasons necessary, dried slides should be frozen at -80°C as soon as possible and stored at this temperature without any freeze-thaw cycles.

### Interindividual differences

Relatively large inter-individual differences, not associated with donors’ age or sex, were observed (data not shown). For example, diffused staining (red present both in cilia and in the cytoplasm) of otherwise stable ciliary proteins was consistently observed in slides from Donor 5, even at optimal storage conditions (**Supplementary Figure S4**). Importantly, ALI culture differentiation of the airway epithelium from Donor 5 enhanced proper ciliary localization of the red signal (**Supplementary Figure S4**), suggesting that epitopes’ stability had been compromised by environmental factors (like in secondary ciliary dyskinesia). Nevertheless, the possibility of a nonspecific cytoplasmic staining even in properly stored slides from healthy donors raises concerns, as it may obscure IF-based PCD diagnostics in cases where pathogenic variants cause diffused cytoplasmic location of the ciliary protein rather than its complete lack from cilia [12, 20]. ALI culture, erasing possible environmental effects, seems to alleviate this problem. However, when IF-based diagnostics is to be made using uncultured airway epithelium samples, it is essential to properly store slides and to take note of patient’s health status at the time of material collection.

### Limitations of the study

Low number of the individuals tested did not allow us to more precisely assess inter-individual differences. Only selected storage conditions and rabbit primary antibodies were tested. We assume that the observed deterioration of the IF staining at some storage conditions reflected targeted epitope instability; moreover, differences in the performance of particular antibodies (even at “optimal” conditions) could reflect antibody polyclonality. Therefore, it is possible that different epitopes of the same ciliary protein have different sensitivity for storage conditions and using antibodies targeting other parts of the protein would improve the IF performance.

## Conclusions

Our study has revealed that proper conditions of the slide storage before IF staining are essential for the reliable PCD diagnosis via specific IF staining. If the microscopic slides are prepared at the diagnostic laboratory, we suggest continuous storage at -80°C or -20°C. If the slides are prepared at a collaborating clinic and shipped, we suggest storage at -20°C until shipment. In any case, the possibility of inter-individual differences should be kept in mind, and IF results indicative of PCD should be always confronted with patient’s clinical history.

## Materials and Methods

### Nasal samples

Nasal specimens from 5 non-PCD donors (average age: 36.4 ± 6.05 yr, without airway infection 4 weeks prior to collection) were collected using a cytological brush and suspended in RPMI1640 medium, as before [21]. Forty µl of cell suspension was pipetted onto a circle of an uncoated cytoslide (309-100-0; Tharmac). Slides dried at RT overnight were stored at different conditions prior to IF analysis **(Table 1)**.The study was conducted according to the guidelines of the Declaration of Helsinki, and approved by the Ethics Committee of Poznan University of Medical Sciences (381/22). A written informed consent was obtained from all participants.

### Immunofluorence microscopy

Samples were fixed and stained as before [21]; for the details see Supplementary material. Each slide was co-stained with one of the primary rabbit antibodies and the mouse AcTub antibody (**Supplementary Table S1**). Images of minimum 30 cells were acquired using tile scan function in LasX software, under, HC PL APO CS2 100x/1.4 OIL objective in the Leica DMi8 confocal microscope. Raw images were exported to tiff images, with each channel saved as separate file.

### Image Analysis

Exported tiff images were analyzed using a custom pipeline created in Cell Profiler software 4.2.7 [22] (main pipeline steps in **Figure 1**). All measurements in CP pipeline are expressed in pixels; to express values in µm, the pixel values were multiplied by the resolution (0.1136 µm/pixel).

Custom R scripts were used to analyze the parameters significantly different (p<0.001) between all samples and to combine all mCilia files for all antibodies and all persons. Initial analysis of the final data was done using pivot tables in Excel, statistics and heatmaps were done using GraphPad Prism. Lack of unified acquisition settings (intensity) of the laser did not allow to analyze the correlation between the intensity of red/green signal depending on the temperature conditions. See **Supplementary Materials and Methods** for further description of the analysis and presented parameters.

## Data Availability

The original raw data presented in the study are available in Zenodo under doi 10.5281/zenodo.15779898.

https://10.5281/zenodo.15463498

## Acknowledgements

The authors would like to thank all the healthy donors for their material. The authors are very grateful Agnieszka Fedoruk-Wyszomirska for the support with confocal imaging. We thank Alicja Rabiasz for critical reading of the manuscript.

## CRediT authorship contribution statement

**Hanna Przystałowska-Macioła:** Investigation, Methodology, Visualization, Writing-review & editing. **Małgorzata Dąbrowska:** Investigation, Methodology, Writing-original draft, Writing-review & editing. **Ewa Ziętkiewicz:** Conceptualization, Supervision, Writing-review & editing. **Zuzanna Bukowy-Bieryłło:** Conceptualization, Methodology, Data curation, Investigation, Validation, Visualization, Writing-original draft, Writing-review & editing.

## Ethics Declaration

The study was conducted according to the guidelines of the Declaration of Helsinki, and approved by the Ethics Committee of Poznan University of Medical Sciences (381/22). A written informed consent was obtained from all participants.

## Data Availability

The original raw data presented in the study are available in Zenodo under doi 10.5281/zenodo.15779898. Preprint of the manuscript has been submitted to Biorxiv under doi

