## Supplementary Data for "Epithelial brushing storage conditions and the reliability of IF staining – implications for primary ciliary dyskinesia diagnostics"

### Supplementary Data Files

### List of contents:

### Supplementary Materials and Methods:

#### Immunofluorescence

Slides were stained as before (ref- TLDA). Briefly, slides were fixed with 4% paraformaldehyde for 15 min and permeabilized with 0,2% Triton X-100/ PBS for 10 min at RT. Slides were blocked with blocking solution (10% BSA and 10% goat serum in 1x PBS) for 1 h at RT. Next, cells were incubated overnight in a moisture chamber at 4˚C with 0,15 ml of primary antibodies diluted in blocking solution (10% BSA and 10% goat serum in 1x PBS). Each slide was co-stained with one of the primary rabbit antibodies and the mouse AcTub antibody (Supplementary Table 1). The slides were observed using Leica DMi8 confocal microscope with LasX software, with HC PL APO CS2 100x/ 1.4 OIL objective using three lasers: 405 nm excitation with 420 to 550 nm detection range (DAPI), 499 nm excitation with 505 to 570 detection (AF488), 590 nm excitation with 600 to 750 nm detection range (AF594); laser intensity settings were adapted for each slide. Images of minimum 30 cells were acquired using tile scan function in LASX software, raw images were exported to tiff files, with each channel saved as separate file.

The pipeline returns eight files (Experiment, Image, Nuclei, Pairs_mTest, Test, mTest, Cilia, mCilia. Experiment and Image files contain the description of the files analyzed, and the analysis parameters. Cilia, Test, and mCilia, mTest files contain the measurements of the objects detected in the green/red channels before or after the object merging steps, respectively. Each measurement file contains the data for all identified objects, and the Children_mTest_Count parameter indicates whether the object has a pair in the other fluorescence channel (0- no pair, >0- paired). The Pairs_mTest file contains the information on the location of objects in the red channel (children) which overlap/pair with objects in the green channel (parents). To assure that the analyzed objects from various channels were present in the same cell, only the paired objects were taken for the parameter analysis.

Out of the 114 parameters measured by the CellProfiler pipeline, 81 parameters were found significantly different between paired objects from positive and negative control samples ($p<0.001$) (**Supplementary Table 2**). The majority of them characterized red channel objects in terms of their area & shape, and intensity (29 and 43 parameters, respectively). The second type of parameters described the red-versus-green channel comparisons, preceded by the definition of paired objects from both channels (seven estimated correlation between the channels, and one measured distance between the centroids of paired objects) (Supplementary Table 1). Out of this group, we have chosen the representatives of each functional group (area/shape, intensity, channel correlation and distance). For the area/shape parameters group, we have chosen object area, as well as two parameters, which characterize the shape of the objects, i.e. area eccentricity and area compactness. Area Eccentricity characterizes how elongated or elliptical an object is, eccentricity values closer to 1 indicate objects that are more elliptical/stretched out, while values closer to 0 indicate more circular shapes. Area Compactness characterizes how "filled" and regular an object is relative to a circle with the same perimeter, compactness values higher than 1 indicate more irregular borders of the object.

Since the laser intensity settings during the image acquisition were not uniform for all slides, in the intensity group, we only selected parameters not directly related to the object intensity (e.g. mean intensity, minimal intensity, etc), but characterize its distribution within the object, e.g. intensity mass displacement. Due to the same reason, in the group of 11 correlation measurements, we have focused on the correlation calculated using the rank-weighted colocalization method (Correlation RWC), as it is a method less sensitive to the differences in overall object intensity levels. Moreover, we have decided to use also the Distance_Centroid_mCilia parameter, which describes the distance between geometric centers of overlapping (paired) objects in the red (tested antibodies) and the green channels (axoneme marker). This measurement is based purely on the spatial locations of the centroids and does not take into account their pixel intensities. This parameter is only calculated when the localization of the object in the red channel (tested antibody) overlaps (pairs) with the object detected in the green channel (anti-Ac-αTub).

**Statistical Analysis**

Distribution of the values in CP profiler results was assessed using D'Agostino & Pearson, Anderson-Darling, Shapiro-Wilk and Kolmogorov-Smirnov distribution tests. Statistical significance of the differences between the storage condition groups and antibodies was assessed in GraphPad using non-parametric Kruskal-Wallis test with Dunn correction for multiple comparisons. Significance of the differences between mean distance to axoneme marker for CCDC39 and other antibodies was assessed in GraphPad Prism using t-test.

### **Supplementary Table S1:** Antibodies used in the study

| Antibody and serial number | Target | Provider, Cat#, RRID | Dilution | Reference |
|---|---|---|---|---|
| | **PRIMARY antibodies** | | | |
| **rabbit anti-DNAH5** | ODA | Atlas Antibodies Cat#HPA037470, RRID:AB_10672348 | 1:800 | [13] |
| **rabbit anti-DNALI1** | IDA | Atlas Antibodies Cat# HPA028305, RRID:AB_10601807 | 1:1000 | [14, 23, 24] |
| **rabbit anti-RSPH9** | RS | Atlas Antibodies Cat#HPA031703, RRID:AB_10602126 | 1:400 | [13, 25, 26] |
| **rabbit anti- RSPH4a** | RS | Atlas Antibodies Cat#HPA031196, RRID:AB_10601612 | 1:400 | [13, 14, 23, 25, 27] |
| **rabbit anti-GAS8** | N-DRC | Atlas Antibodies Cat#HPA041311, RRID:AB_2677402 | 1:600 | [13, 28] |
| **rabbit anti-CCDC39** | MR | Atlas Antibodies Cat#HPA035364, RRID:AB_10601095 | 1:500 | [17] |
| **rabbit anti-CCDC40** | MR | Atlas Antibodies Cat#HPA022974, RRID:AB_1848791 | 1:1600 | [24] |
| **rabbit anti- Spef2** | CP | Atlas Antibodies Cat#HPA039606, RRID:AB_10671044 | 1:300 | [27, 29, 30] |
| **mouse** anti-acetylated α- tubulin (**Ac-αTub**) | MTs, axonemal marker | Sigma-Aldrich Cat# T6793, RRID:AB_477585 | 1:10,000 | [23, 31] |
| | **SECONDARY antibodies** | | | |
| AlexaFluor 594 goat anti-rabbit IgG (H+L) | Tested antibodies (red) | A11037, Invitrogen | 1:2,500 | [15, 21] |
| AlexaFluor 488 goat anti-mouse IgG (H+L) | **Ac-**αTub (green) | A11029, Invitrogen | 1:2,500 | [15, 21] |

**Supplementary Table S2:** Correlations between the storage temperatures and analyzed image/ object parameters.

Names of the columns contain the measurement group (AreaShape, Intensity, Correlation etc), followed by the name of the parameter measured (e.g. Area, Eccentricity etc). Names of the columns containing only the measurement group and the parameter name refer to measurements performed on the green channel. Column names with suffix "Mean_mTest" refer to the measurements done on the red channel. Names of the parameters (column 1) chosen for further analyses are shown in bold. P-values (column 3) lower than 0.001 are shown in bold.

| Column | Rho | P_value |
|---|---|---|
| **Mean_mTest_Correlation_RWC_RED_GREEN** | -0.115661134163905 | **1.4731882347589e-67** |
| Mean_mTest_Correlation_Manders_RED_GREEN | -0.111155174785092 | **1.66073648270303e-62** |
| Mean_mTest_Intensity_MassDisplacement_GREEN | 0.110559925050577 | **7.45421277797541e-62** |
| Mean_mTest_Intensity_LowerQuartileIntensity_GREEN | -0.095947036415811 | **5.85253404288507e-47** |
| Intensity_MeanIntensityEdge_RED | 0.0873069756130233 | **3.74086142571518e-39** |
| Intensity_MinIntensity_RED | 0.0852235041321796 | **2.21063353004736e-37** |
| Intensity_MinIntensityEdge_RED | 0.0851610998501053 | **2.49411337507631e-37** |
| Intensity_MaxIntensityEdge_RED | 0.083886178816177 | **2.87750123785879e-36** |
| Mean_mTest_Intensity_MedianIntensity_GREEN | -0.0838493629037626 | **3.08636498961121e-36** |
| Intensity_StdIntensityEdge_RED | 0.0817099968279706 | **1.71734278255422e-34** |
| Mean_mTest_Intensity_MaxIntensity_RED | 0.0814502192120748 | **2.77794435987991e-34** |
| Mean_mTest_Intensity_IntegratedIntensityEdge_RED | 0.0797727878882791 | **5.97584730214557e-33** |
| **Mean_mTest_AreaShape_Eccentricity** | -0.0745026468670747 | **6.07065770029549e-29** |
| **Mean_mTest_AreaShape_Compactness** | 0.0739983777426539 | **1.42011102755787e-28** |
| Mean_mTest_AreaShape_FormFactor | -0.073998375987077 | **1.42011521504808e-28** |
| Intensity_LowerQuartileIntensity_RED | 0.0734700337700935 | **3.43837580785071e-28** |
| **Mean_mTest_Intensity_MassDisplacement_RED** | 0.0709328993172888 | **2.19970517734353e-26** |
| Intensity_MedianIntensity_RED | 0.0705986203437883 | **3.76368199691447e-26** |
| Mean_mTest_AreaShape_MinorAxisLength | 0.068394778517376 | **1.21901537395203e-24** |
| Intensity_MeanIntensity_RED | 0.0682695941226386 | **1.48039695911839e-24** |
| Mean_mTest_AreaShape_MinFeretDiameter | 0.0681015125610288 | **1.92051257338006e-24** |
| AreaShape_Area | -0.0675331148691324 | **4.60927906596462e-24** |
| Mean_mTest_Correlation_Overlap_GREEN_RED | -0.0667691266729029 | **1.47812779039696e-23** |
| Intensity_MinIntensityEdge_GREEN | 0.0663331307321573 | **2.85738363414516e-23** |
| Mean_mTest_AreaShape_MaximumRadius | 0.0648248365524599 | **2.70358674180827e-22** |
| Mean_mTest_Intensity_MeanIntensity_GREEN | -0.0647380814496224 | **3.07186379928636e-22** |
| Intensity_UpperQuartileIntensity_RED | 0.0643785951368104 | **5.2051799925484e-22** |
| Mean_mTest_AreaShape_Perimeter | 0.0641630917112213 | **7.13069689497273e-22** |
| Mean_mTest_Intensity_IntegratedIntensity_RED | 0.0633740560622559 | **2.23750212610428e-21** |
| **Mean_mTest_Distance_Centroid_mCilia** | 0.0626689364997743 | **6.14390647972798e-21** |
| AreaShape_MajorAxisLength | -0.0597616850928501 | **3.51598881603417e-19** |
| Intensity_MinIntensity_GREEN | 0.0579942240969737 | **3.7533998249285e-18** |
| Mean_mTest_AreaShape_EulerNumber | -0.0576423867542222 | **5.96372981612822e-18** |
| AreaShape_MinorAxisLength | -0.0562887158860364 | **3.45154428691022e-17** |
| AreaShape_MaxFeretDiameter | -0.0548696608807477 | **2.08099471003424e-16** |
| Mean_mTest_AreaShape_BoundingBoxMinimum_X | -0.0548510547286008 | **2.12996224180127e-16** |
| Intensity_MaxIntensity_RED | 0.0546908662299549 | **2.60133675927785e-16** |

| | | |
|---|---|---|
| Mean_mTest_Intensity_UpperQuartileIntensity_GREEN | -0.052940785557876 | **2.22635104016063e-15** |
| Mean_mTest_AreaShape_ConvexArea | 0.0528858975100879 | **2.37879766369995e-15** |
| Mean_mTest_Intensity_MinIntensityEdge_RED | 0.0527225505351725 | **2.89591991616328e-15** |
| AreaShape_MinFeretDiameter | -0.052585927370506 | **3.41225304014872e-15** |
| Mean_mTest_Intensity_MinIntensityEdge_GREEN | -0.0521190663563624 | **5.95891986688447e-15** |
| **Mean_mTest_AreaShape_Area** | 0.05205459139912 | **6.43339864513345e-15** |
| Mean_mTest_AreaShape_EquivalentDiameter | 0.0520545399707035 | **6.43379157048685e-15** |
| Mean_mTest_AreaShape_BoundingBoxArea | 0.0509127093002062 | **2.46075771011882e-14** |
| Mean_mTest_Intensity_MeanIntensityEdge_GREEN | -0.0502886572747398 | **5.06015969494861e-14** |
| AreaShape_Perimeter | -0.0499470451551088 | **7.48108827749426e-14** |
| Mean_mTest_Intensity_MinIntensity_GREEN | -0.0495536753389163 | **1.16976361187347e-13** |
| AreaShape_EulerNumber | 0.0485424087238789 | **3.63352377033922e-13** |
| Mean_mTest_Intensity_StdIntensityEdge_RED | -0.0470784320080157 | **1.80081495553667e-12** |
| Mean_mTest_AreaShape_BoundingBoxMinimum_Y | -0.046183348807341 | **4.68089098183343e-12** |
| Intensity_UpperQuartileIntensity_GREEN | 0.0442933552846822 | **3.31925637184409e-11** |
| Mean_mTest_Location_MaxIntensity_X_GREEN | -0.0435784351882528 | **6.82189349171669e-11** |
| Mean_mTest_AreaShape_Center_X | -0.0433915942188394 | **8.21983837019178e-11** |
| Intensity_MADIntensity_RED | 0.0433576427917249 | **8.50234174526413e-11** |
| Mean_mTest_Location_CenterMassIntensity_X_RED | -0.0432470193119049 | **9.49026271564208e-11** |
| Mean_mTest_Location_MaxIntensity_X_RED | -0.0430592735917971 | **1.14296222797793e-10** |
| Mean_mTest_Intensity_MinIntensity_RED | 0.0430049098018489 | **1.20601289281675e-10** |
| Mean_mTest_Location_CenterMassIntensity_X_GREEN | -0.0426035200298296 | **1.78919578623249e-10** |
| Intensity_MeanIntensity_GREEN | 0.0417880900271573 | **3.94386409917333e-10** |
| Mean_mTest_AreaShape_Solidity | -0.0417533884733877 | **4.07745217281211e-10** |
| Mean_mTest_Correlation_RWC_GREEN_RED | -0.0414159693658734 | **5.62933921704592e-10** |
| Mean_mTest_Intensity_MADIntensity_GREEN | -0.0412655604906338 | **6.4944767114122e-10** |
| Mean_mTest_AreaShape_MaxFeretDiameter | 0.0406750110791186 | **1.13298733305405e-09** |
| Mean_mTest_Correlation_Costes_RED_GREEN | -0.0385137101883461 | **8.13506572571191e-09** |
| Intensity_MedianIntensity_GREEN | 0.0381943449244061 | **1.07918100973505e-08** |
| Correlation_Overlap_GREEN_RED | -0.0373095094140293 | **2.33384485994372e-08** |
| Mean_mTest_Intensity_StdIntensity_GREEN | 0.0371274733314242 | **2.72937650226708e-08** |
| Mean_mTest_Correlation_Costes_GREEN_RED | -0.0367984971700827 | **3.6152585605092e-08** |
| Mean_mTest_AreaShape_MeanRadius | 0.0354448544818111 | **1.12108654845955e-07** |
| Correlation_RWC_RED_GREEN | -0.0353938965663508 | **1.1689672588802e-07** |
| Intensity_StdIntensity_GREEN | 0.0351319216284991 | **1.44807549248052e-07** |
| Mean_mTest_Location_CenterMassIntensity_Y_GREEN | -0.0349418979122752 | **1.68979509999105e-07** |
| Intensity_MassDisplacement_RED | -0.03485817340441 | **1.80827572656922e-07** |
| Mean_mTest_Location_MaxIntensity_Y_GREEN | -0.0343282580007987 | **2.76694482502182e-07** |
| Mean_mTest_Location_CenterMassIntensity_Y_RED | -0.0338470463007074 | **4.05001749894641e-07** |
| Mean_mTest_AreaShape_Center_Y | -0.0338181061082394 | **4.14321741439672e-07** |
| Intensity_MassDisplacement_GREEN | -0.0330454271369436 | **7.554798045844e-07** |
| Intensity_IntegratedIntensity_GREEN | -0.0326780597590011 | **1.00065336177349e-06** |
| Mean_mTest_Location_MaxIntensity_Y_RED | -0.0325549017497455 | **1.09880113624653e-06** |
| Intensity_LowerQuartileIntensity_GREEN | 0.0322175373480849 | **1.41742124739326e-06** |
| Mean_mTest_AreaShape_BoundingBoxMaximum_X | -0.0318255701059463 | **1.89945944178851e-06** |
| Intensity_IntegratedIntensityEdge_RED | 0.0290745932645528 | **1.34979854771562e-05** |
| Mean_mTest_Intensity_StdIntensity_RED | 0.0290049336438959 | **1.41551084515568e-05** |

| | | |
|---|---|---|
| Intensity_MeanIntensityEdge_GREEN | 0.0288940343984262 | **1.52646045089015e-05** |
| Intensity_StdIntensity_RED | 0.0288407960591754 | **1.58262281187249e-05** |
| Intensity_MADIntensity_GREEN | 0.0276190266137396 | **3.56630114366653e-05** |
| AreaShape_Extent | -0.0271019179725907 | **4.98155437207045e-05** |
| Mean_mTest_AreaShape_MajorAxisLength | 0.0268328229207534 | **5.91442660797361e-05** |
| Mean_mTest_Correlation_Correlation_GREEN_RED | -0.0242258164603437 | **0.000287984490430112** |
| Correlation_Correlation_GREEN_RED | -0.0239038606181855 | **0.000346665205593237** |
| Mean_mTest_Intensity_MaxIntensity_GREEN | 0.0233843661033745 | **0.000465432217230539** |
| Intensity_MaxIntensity_GREEN | 0.0229615480818657 | **0.000589062772197361** |
| Mean_mTest_AreaShape_BoundingBoxMaximum_Y | -0.0227881577322848 | **0.000648098259255224** |
| Intensity_IntegratedIntensityEdge_GREEN | -0.022195163565802 | **0.000894165885623938** |
| Intensity_StdIntensityEdge_GREEN | -0.0221531136936755 | **0.000914550465971032** |
| Mean_mTest_Intensity_MeanIntensityEdge_RED | 0.0209522315742683 | 0.00171384826553063 |
| AreaShape_Solidity | -0.0192271807084375 | 0.00400746437963143 |
| Mean_mTest_AreaShape_Extent | -0.0184735492135954 | 0.00569639697011823 |
| Mean_mTest_AreaShape_MedianRadius | 0.0169250697204568 | 0.0113096471185149 |
| Mean_mTest_Intensity_MedianIntensity_RED | -0.0158943917498985 | 0.0173732006003841 |
| Mean_mTest_Correlation_K_RED_GREEN | -0.0158744478057185 | 0.0175143787030025 |
| Mean_mTest_Intensity_MaxIntensityEdge_GREEN | 0.0152033368700986 | 0.0228891846375894 |
| AreaShape_Compactness | -0.0151555623294804 | 0.0233213676682738 |
| AreaShape_FormFactor | 0.0151555623294804 | 0.0233213676682738 |
| Mean_mTest_Intensity_IntegratedIntensity_GREEN | 0.0140226254031119 | 0.0358557148246747 |
| Intensity_IntegratedIntensity_RED | -0.0129660589300417 | 0.0523280229893498 |
| Mean_mTest_Intensity_MaxIntensityEdge_RED | 0.0118833783332502 | 0.0753405397976296 |
| Mean_mTest_Correlation_K_GREEN_RED | 0.0106770412020535 | 0.110078022545708 |
| Mean_mTest_Intensity_StdIntensityEdge_GREEN | 0.0099257964788003 | 0.137434881265135 |
| Mean_mTest_Correlation_Manders_GREEN_RED | 0.0085077506460563 | 0.202950547026143 |
| Mean_mTest_Intensity_LowerQuartileIntensity_RED | -0.00790921666355896 | 0.236564971341632 |
| Mean_mTest_Intensity_IntegratedIntensityEdge_GREEN | 0.00757717244794935 | 0.256826272244064 |
| AreaShape_Eccentricity | -0.0073334098797523 | 0.272447220805256 |
| Mean_mTest_Intensity_MADIntensity_RED | -0.00477117033372176 | 0.475227315149796 |
| Mean_mTest_AreaShape_Orientation | -0.00388568121761845 | 0.560912743905971 |
| Mean_mTest_Intensity_UpperQuartileIntensity_RED | -0.00354038895209794 | 0.596241241856369 |
| Intensity_MaxIntensityEdge_GREEN | -0.00290160791932087 | 0.664128324904755 |
| Correlation_RWC_GREEN_RED | 0.000516425948932618 | 0.938399150872506 |
| Mean_mTest_Intensity_MeanIntensity_RED | -0.000252977733007192 | 0.969801225317864 |

**Supplementary Table S3:** *Overall performance of analyzed antibodies calculated on storage conditions excluding prolonged RT storage.*

For each of the analyzed parameters, antibodies were ranked according to the mean parameter's value for all storage conditions (see rightmost columns in the heatmaps). The overall performance rank was calculated as the mean from all parameters. Colors of the bars below the table indicate the performance (green – very good, yellow – average, red – poor). Calculations were made for green objects pairing with only 1 red object.

| | Antibodies: | | | | | | | |
|---|---|---|---|---|---|---|---|---|
| | RSPH4A | DNAH5 | GAS8 | SPEF2 | RSPH9 | CCDC40 | DNALI1 | CCDC39 |
| Mean_mTest_Correlation_RWC_RED_GREEN | 1 | 2 | 7 | 4 | 6 | 5 | 3 | 8 |
| Mean_mTest_AreaShape_Compactness | 1 | 3 | 2 | 5 | 4 | 6 | 7 | 8 |
| Mean_ mTest_AreaShape_Area | 1 | 2 | 3 | 5 | 4 | 6 | 8 | 7 |
| Mean_mTest_Area_Eccentricity | 1 | 3 | 6 | 2 | 7 | 5 | 4 | 8 |
| Mean_mTest_Intensity_Mass_Displacement | 1 | 2 | 5 | 4 | 3 | 6 | 7 | 8 |
| Mean_mTest_Distance_Centroid_mCilia | 4 | 1 | 2 | 5 | 3 | 6 | 7 | 8 |
| | 1,5 | 2,2 | 4,2 | 4,2 | 4,5 | 5,7 | 6,0 | 7,8 |

**Supplementary Figure S1**. Violin plots for the Distance_Centroid_mCilia_um values for various tested antibodies under different storage conditions.

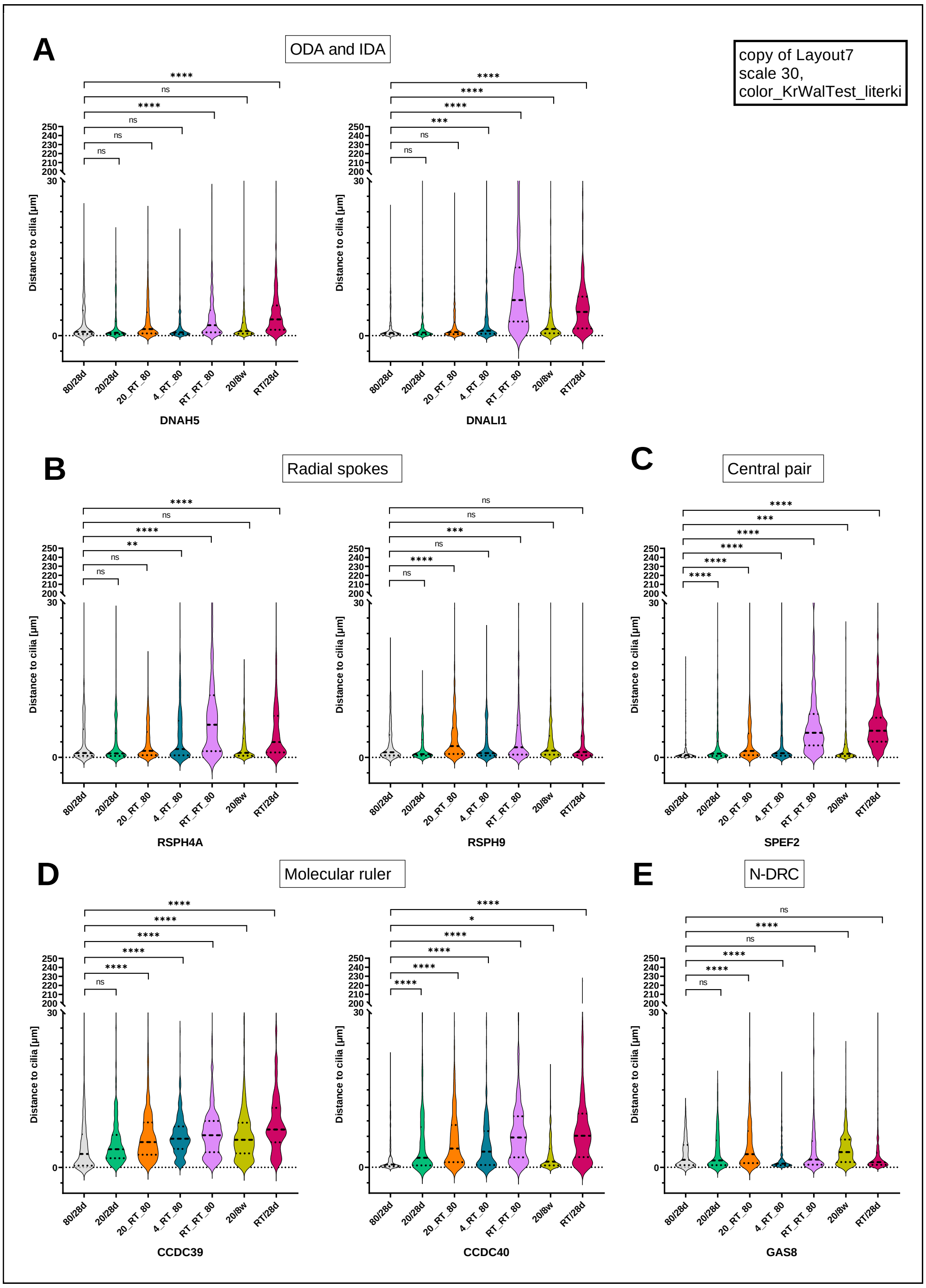

*Figure 4. Distribution of the distances between the centroids of paired red and green objects (Distance_Centroid_mCilia). A: ODA and IDA proteins; B: RS proteins; C:CP protein; D: MR proteins; E: N-DRC proteins. Thick dotted lines indicate median values, thin dotted lines indicate first and third quartile of the distance. Pairwise-comparisons between the centroid distance at the optimal storage conditions (-80oC for 28d) and other tested conditions were done using Kruskal-Wallis test with Dunn correction for multiple testing. *** indicates* $p < 0.001$*, * indicates* $p < 0.01$, $p < 0.05$, ns = not significant*).*

Violin plots displaying the distribution of the distances for each antibody at all tested storage conditions are shown in Supplementary Data **Supplementary Figure 3)**. Violin plots for the optimal storage condition (-80oC/28d; "positive control") for the analyzed antibodies revealed a weak dispersion of the distance values, the majority of them concentrated around the median, which was itself localized close to the 0 value. As expected, the "negative control" (RT/28d) was significantly different, with much higher median and range of the distance values. A comparably deleterious effect was observed for slides stored at RT for 14 days, followed by 14 days at -80oC (**Supplementary Figure 3**).

### **Supplementary Figure S2** Influence of the fluorochrome color on the results of Cell Profiler analysis for various channels.

Slides stored at -80oC_28d and RT_28d were co-stained for DNALI1 and axoneme marker, using secondary antibodies associated with fluorochromes of various colors (green line– Alexa Fluor 488, red line– Alexa Fluor 594). While the fluorochrome switch resulted in the switch of the corresponding values, the relative differences between the best and worst storage conditions remained similar.

Left panel: results for DNALI1 antibody; right panel: results for axoneme marker. The color of the line indicates the color of the fluorochrome used in the staining/ channel color. Numbers on the X axis indicate the storage conditions: T1 (-80C_28d), T2 (RT_28d) or the mean values between the conditions (the mean is shown for informative purposes only).

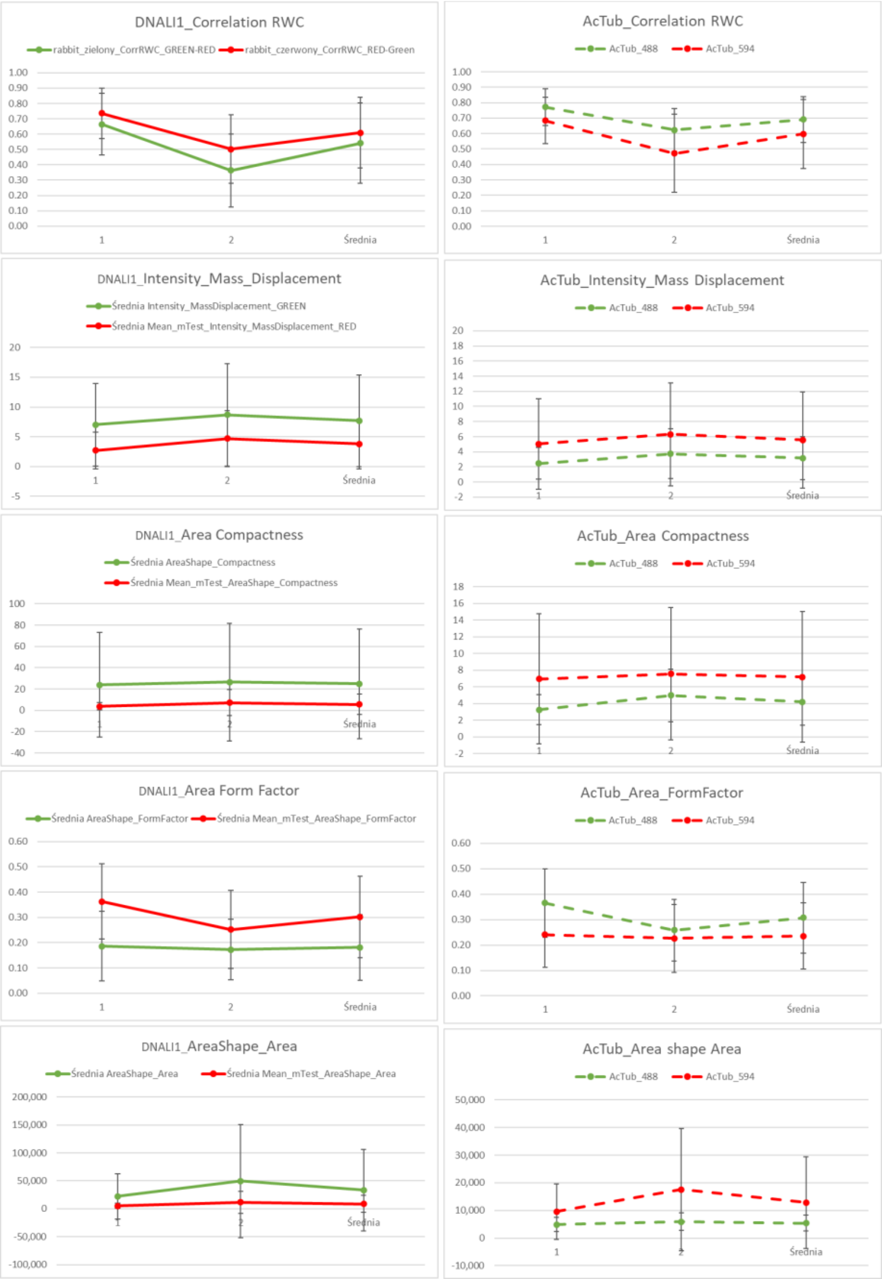

**Supplementary Figure S3:** Comparison of the values of the Distance_Centroid_mCilia for individual donors.

Data are expressed in um, points display the median values for individual donors +/- median absolute deviation (MAD).

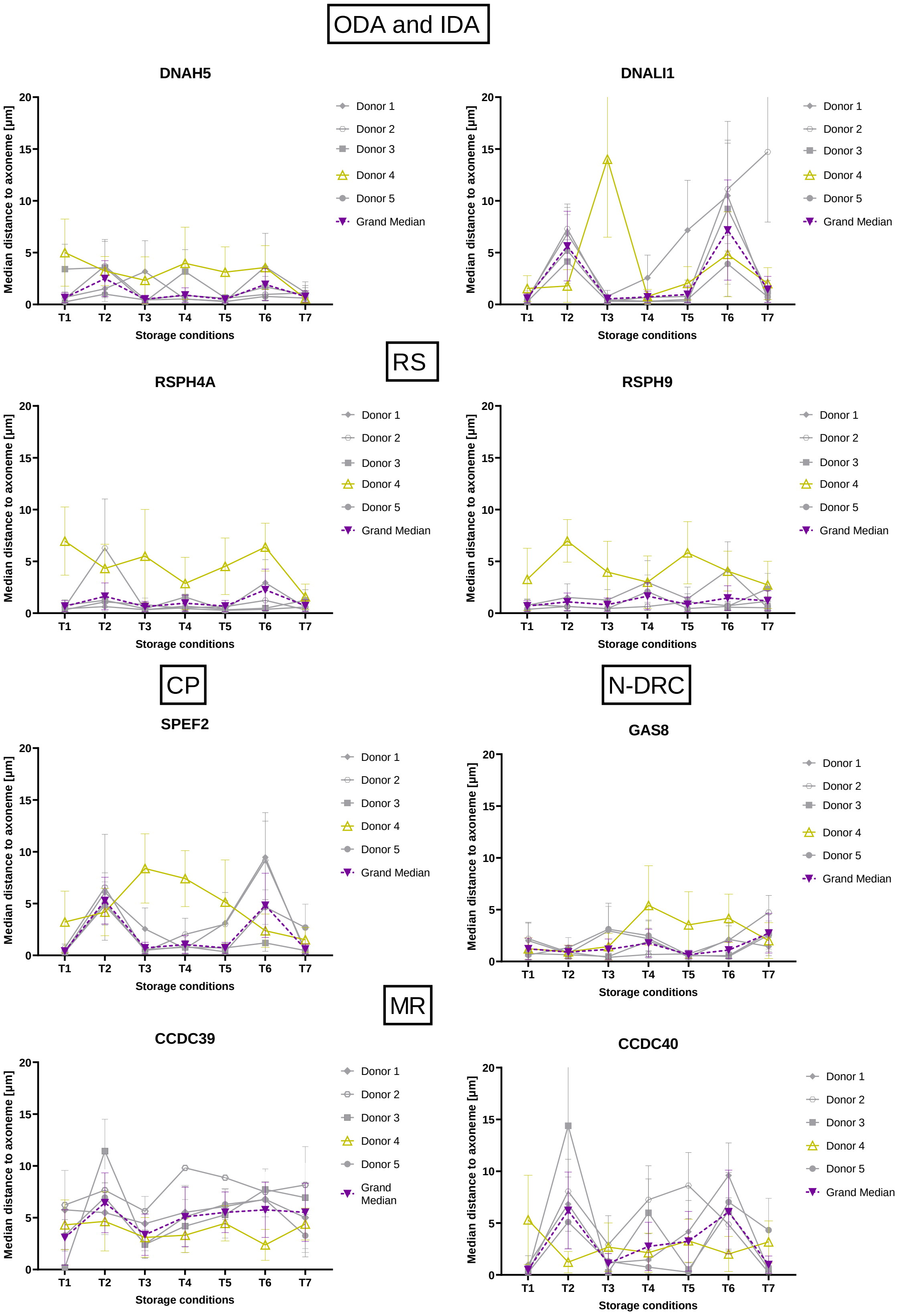

**Supplementary Figure S4:** IF staining of donor 5 nasal cells before and after in vitro differentiation/ALI culture.

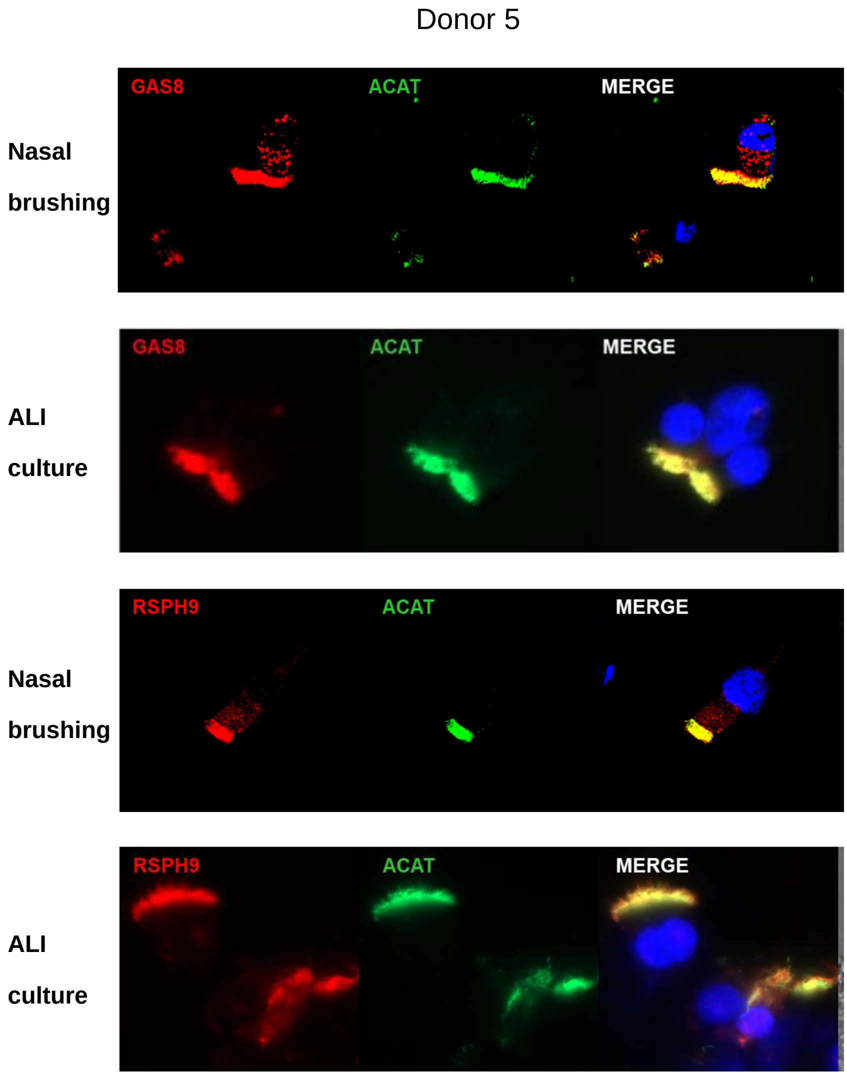
